# PPP-COMPUTE: A Pandemic Pharmacology Platform for COMPUTational Evaluation of Anti-infectives in Pandemic Preparedness

**DOI:** 10.64898/2026.07.30.26359333

**Authors:** Qinhao Wu, Chenyu Wang, Wisse van Os, Xuanlin Liu, Jiahang Su, Anne-Grete Märtson, J. G. Coen van Hasselt, Linda B.S. Aulin, Tingjie Guo

## Abstract

A global pandemic requires accelerated strategies to mitigate public healthcare risks and preserve societal stability, either by repurposing existing drugs or by rapidly developing novel drug candidates. Computational platforms have accelerated candidate identification for both routes. A critical gap is the translation from *in vitro* potency to predicted clinical efficacy, which determines whether a prioritized candidate can achieve therapeutic effect under realistic dosing. To address this, we developed the Pandemic Pharmacology Platform for COMPUTational Evaluation of anti-infectives (PPP-COMPUTE), a comprehensive pharmacokinetic/pharmacodynamic (PK/PD) simulation platform designed to support evaluation and prioritization of drug candidates and clinical trial design during a pandemic. PPP-COMPUTE integrates experimentally derived anti-infective potency data (e.g., *EC*_50_) with PK/PD modeling to evaluate whether clinical dosing regimens can achieve sufficient exposure for therapeutic efficacy in patients. The platform incorporates mechanism-based dynamic models with a focus on viral pathogens to simulate time-dependent viral load trajectories under various treatment scenarios, enabling quantitative assessment of antiviral response and optimization of dosing strategies. Probability of target attainment analyses further support evaluation of regimen feasibility against predefined pharmacological targets. The clinical trial module generates simulated virological endpoints to evaluate candidate clinical study designs. PPP-COMPUTE is an accessible and quantitative framework that links PK and preclinical anti-infective potency data with predicted clinical benefit, thereby supporting rapid drug evaluation during future pandemics.

## 1. Introduction

Pandemics pose a substantial threat to public health, economies, and social stability, often requiring urgent and coordinated international responses. The COVID-19 pandemic exemplified the scale and duration such crises can reach, which resulted in significant mortality and socioeconomic disruption worldwide [1]. Since the COVID-19 pandemic, there has been increased attention to enhance preparedness for future outbreaks of emerging pathogens [2, 3]. During rapidly evolving global health emergencies, developing therapeutic interventions in a timely fashion is critical. Alongside the development of new drugs, drug repurposing represents another strategy, in which existing anti-infective agents approved for other infectious diseases are evaluated against an emerging pathogen. This is a practical approach to accelerate treatment by leveraging compounds with established pharmacological, clinical, and safety data [4, 5, 6].

Numerous computational platforms have been developed to accelerate drug repurposing [6]. For instance, CoVex [7] integrates virus-host-drug interactome data to support target identification, while network-based approaches such as SAveRUNNER [8], Dr-COVID [9, 10], and graph convolutional models [11] apply graph and deep-learning methods to prioritize candidate drugs. More recently, unsupervised machine-learning frameworks have been used to cluster heterogeneous drug data from the literature for candidate selection [12]. These approaches are valuable for prioritizing candidates based on *in vitro* potency, but their focus is largely on drug–target interactions (e.g., docking and pathway analysis) and predicted bioactivity. However, there is limited attention to whether that activity translates into a therapeutic response under realistic drug exposures [13, 14]. As a result, *in vitro* potency alone cannot confirm whether a prioritized compound will be effective at clinically achievable exposures. This leaves a gap between predicted bioactivity and the expected therapeutic effect.

Pharmacokinetic (PK) and pharmacodynamic (PD) modeling addresses this gap by providing a mechanism-based, quantitative link between drug exposure and therapeutic effect. To determine whether a candidate drug can be clinically effective, both the drug exposure achieved under a given regimen (i.e., PK) and the corresponding therapeutic effect (i.e., PD) must be predicted. PK/PD models link the concentration-time course of the drug to treatment effect, such as pathogen killing, enabling evaluation of whether adequate treatment effect can be achieved at clinically relevant doses. PK/PD models can integrate preclinical efficacy data with human PK to simulate treatment response and support clinical translation. This approach supports quantitative exposure-response analysis, dose optimization, and clinical trial design, and can be used to predict the clinical benefit before conducting large-scale clinical studies [15].

In this study, we developed the Pandemic Pharmacology Platform for COMPUTational Evaluation of anti-infectives (PPP-COMPUTE), a comprehensive PK/PD simulation platform designed to support rapid drug evaluation during a pandemic. The platform supports both repurposing of approved drugs and development of novel candidates. Here we provide a detailed technical overview of its components and workflow. The platform integrates experimentally derived antiviral potency data, such as the *EC*_50_ obtained from *in vitro* studies, with PK modeling to evaluate whether clinically relevant dosing regimens can attain sufficient treatment effect in patients. In addition, the platform incorporates mechanistic viral dynamic models for simulation of time-dependent viral load trajectories across different dosing regimens. This provides a quantitative basis for comparing treatment strategies and informing key aspects of clinical trial design. Together, these components bridge preclinical potency findings with predicted clinical efficacy.

## 2. Methods

PPP-COMPUTE comprises three core modules: the PK Module, the PD Module, and the clinical trial Module, shown in Figure 1. The platform facilitates PK and PD analyses to support anti-infective drug development, with a focus on antivirals, and clinical trial design during emerging pandemics. Probability of target attainment (PTA) analyses based on PK/PD simulations evaluate whether approved drugs and dosing regimens can achieve effective exposures sufficient in relation to half maximal effective concentration (*EC*_50_) against a pathogen. Additionally, PK/PD simulations can also characterize the effects of candidate therapies on viral clearance and generate virological endpoints to inform clinical trial design. The following subsections describe the core modules, the user interface (UI), and the associated curated database, PPP-db.

**Figure 1:**
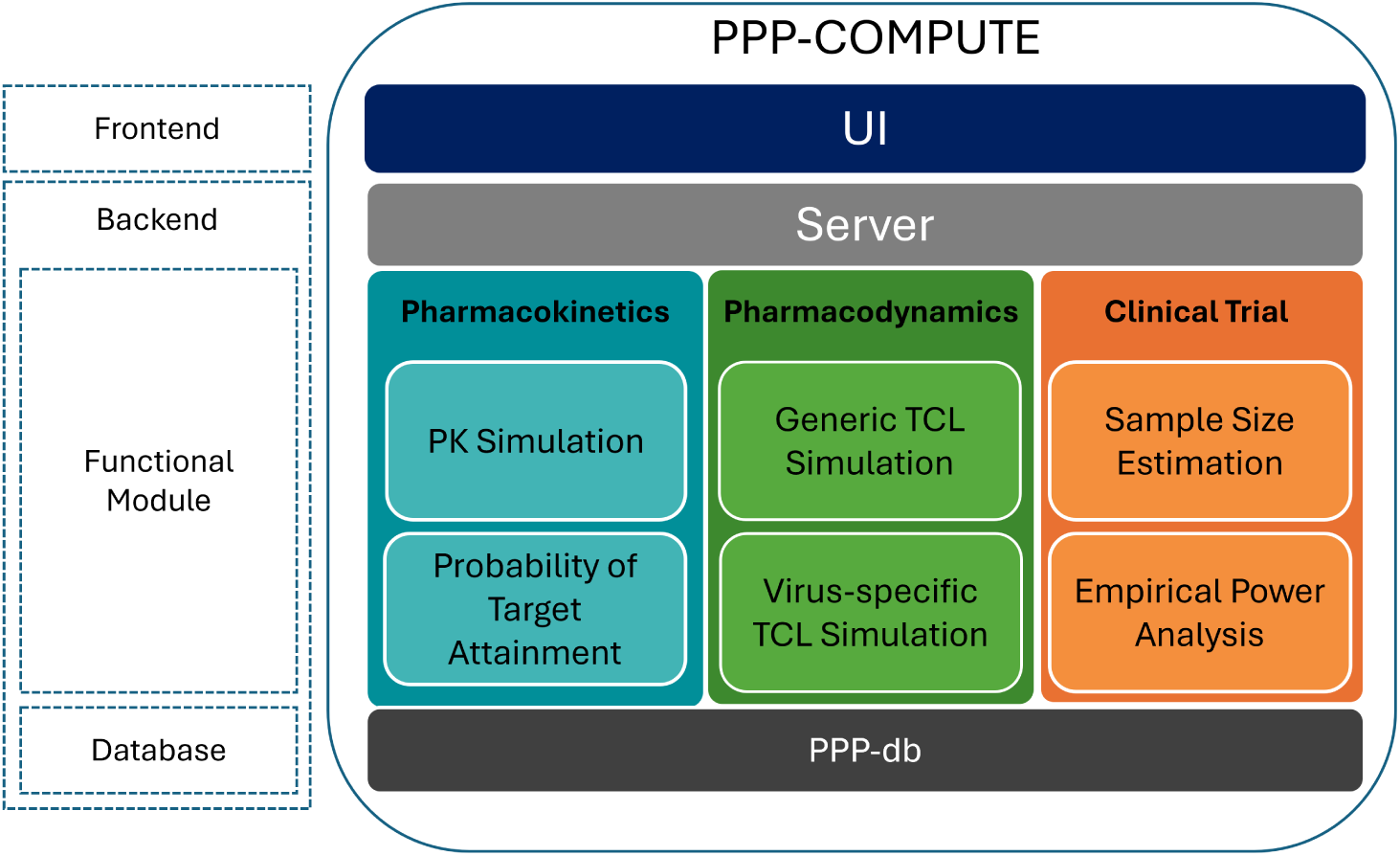
Overview of the PPP-COMPUTE platform.

### 2.1. Pharmacokinetics module

The PK module can be used to simulate drug concentration–time profiles in plasma and selected highly perfused organs including lung, liver, and kidney. Drug concentrations in organs are empirically estimated using tissue partition coefficients without accounting for mass balance. Users can evaluate drug exposure using either non-compartmental analysis-(NCA) derived PK parameters collected from DrugBank or published population pharmacokinetic (PopPK) models. For new compounds, PK parameters can be predicted from Simplified Molecular Input Line Entry System (SMILES) strings using a previously developed Quantitative Structure–Property Relationship (QSPR) model [16, 17]. Specifically, the PK parameters derived from NCAs or predicted from QSPR are used to simulate the drug concentration in plasma via a one-compartment model. This choice is made due to its ability to support efficient and consistent simulation of drug exposure in data-limited settings without requiring additional mechanistic assumptions.

Simulated concentration–time profiles incorporate inter-individual variability (IIV) and enable calculation of the PTA, defined as the proportion of simulated individuals achieving unbound concentrations above the *in vitro EC*_50_ or *EC*_90_ at the target site. PTA analysis provides a rapid assessment of whether a dosing regimen can achieve sufficient drug exposure in plasma or at the target site.

### 2.2. Pharmacodynamics module

The PD module can be used to simulate viral load trajectories within host cells using mechanistic viral dynamic models. Both generic and virus-specific target cell limited (TCL) models with IIV are implemented. By integrating PK models with viral dynamic models, the module enables simulation of treatment effects for compounds available in the database or provided by the user, allowing evaluation of their impact on viral load under different dosing regimens. These simulations support quantitative assessment of antiviral efficacy and disease progression.

### 2.3. Clinical trial module

The clinical trial module supports clinical trial design informed by PK/PD simulations. By linking simulated drug exposure to virological endpoints, such as viral load and the area under the viral load curve (AUC), the module quantifies expected treatment effects under different dosing regimens and study designs. Based on these simulations, it is possible to conduct statistical power analyses and evaluate the sample sizes required to detect clinically meaningful treatment effects.

### 2.4. User interface

PPP-COMPUTE consists of a web-based user interface (UI) connected to a backend infrastructure that includes a server, functional modules, and a curated database (PPP-db) (Figure 1). Access to the main functionalities is organized through sidebar navigation, allowing users to interact with the PK/PD simulation and clinical trial design tools through dedicated interfaces (Supplementary Materials Figures S1 and S2).

The “PK/PD Simulation” interface is organized into five sections: *Virus Selection*, *Drug and Pharmacokinetic Parameters*, *Patient Settings*, *Treatment*, and *Pharmacodynamics*. These sections guide users through a structured workflow in which available options are progressively refined based on previous selections. The *Drug and Pharmacokinetic Parameters* section provides access to key PK functionalities, including prediction of PK parameters for new compounds, selection of approved drug parameters, and implementation of published PopPK models. The *Patient Settings* and *Treatment* sections allow users to specify patient characteristics, including covariates from selected population models, and to define dosing regimens. The *Pharmacodynamics* section allows selection of PK/PD targets and viral dynamic models with user-defined parameters for PK/PD simulations. The “Clinical Trial Design” interface builds on the PK/PD simulation and provides the inputs required for sample size estimation and power analysis (Supplementary Materials Figure S2).

### 2.5. The PPP database (PPP-db)

PPP-db is a core component of PPP-COMPUTE and contains drug- and virus-centered information required for PK/PD simulations and clinical trial design. The database is organized around approved drugs and viral pathogens and contains information on drug physicochemical properties, PK parameters, a PopPK model library, dosing regimens, and antiviral bioassay data. It comprises nine tables where *Approved_Drug* is the central table. It is linked with eight satellite tables that capture distinct pharmacological characteristics, including PK parameters, tissue distribution, clinical dosing regimens, and antiviral bioassay activity (Figure 2). Further details of PPP-db are provided in the Supplementary Materials.

**Figure 2:**
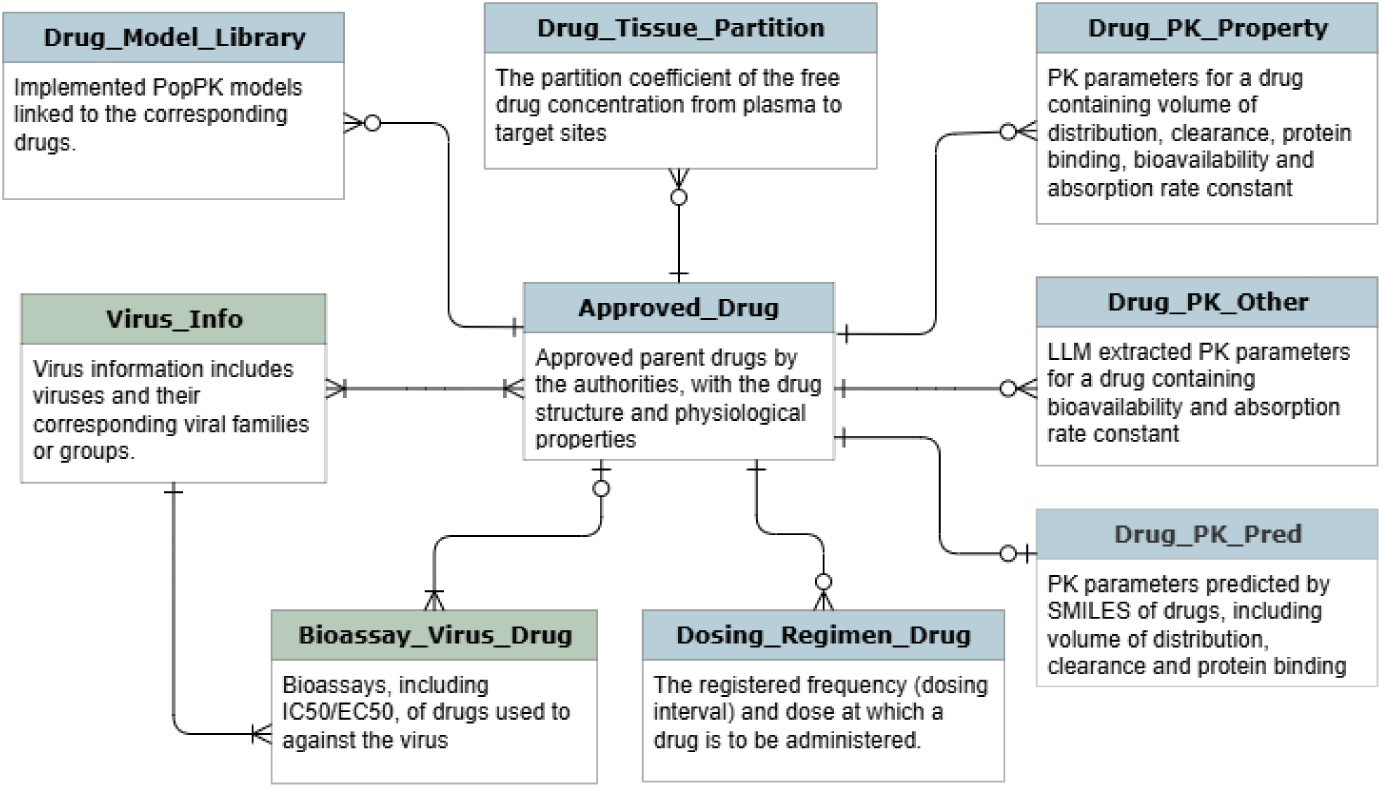
The database schema overview of PPP-db. Tables are connected using crow’s foot notation [31], in which a dash next to a table denotes “one”, a circle denotes “zero”, and a fork denotes “many”. Table titles are color-coded: blue corresponds to the PK module and green corresponds to the PD module.

PPP-db was curated from multiple open-source databases, including ChEMBL (Version 34) [18, 19], DrugBank (Version 5.1.12) [20], DailyMed [21], openFDA [22], Drugs.com [23], Medscape [24], and Farmacotherapeutisch Kompas [25], as well as published studies [26, 27, 28] and previously published PopPK models (see Supplemental Material). The database was constructed in accordance with the FAIR (Findable, Accessible, Interoperable, and Reusable) principles to facilitate integration into PPP-COMPUTE. Detailed extraction procedures are provided in the Supplementary Material.

The central component of the PPP-db is the Approved_Drug table, which contains the list of approved compounds available for PK/PD simulations. Each drug entry is linked to virus information and antiviral bioassay data through the Virus_Info and Bioassay_Virus_Drug tables, which provide key antiviral activity measures including *IC*_50_ and *EC*_50_ values used in the PD and CT modules. PK-related information is organized in dedicated tables supporting the PK module. The Drug_Model_Library, Drug_Tissue_ Partition, and Dosing_Regimen_Drug tables contain published PopPK models, estimated tissue partition coefficients, and clinically approved dosing regimens, respectively. Additional PK parameters are stored in Drug_PK_ Property, Drug_PK_Pred, and Drug_PK_Other. The Drug_PK_Property and Drug_PK_Other tables include parameters extracted from DrugBank, such as clearance and volume of distribution, as well as oral absorption parameters including bioavailability and absorption rate constants. The Drug_PK_Pred table contains QSPR model predictions of intravenous PK parameters for compounds lacking clinical PK information.

PK parameters were collected using multiple complementary approaches. Volume of distribution (Vd), clearance (CL), and free fraction (fu) were initially extracted from DrugBank using rule-based parsing methods and combined with curated datasets from Lombardo *et al.* [27] and Mamada *et al.* [28], resulting in coverage of 1276 compounds for Vd and CL and 739 compounds for fu. To further expand parameter coverage, large language model (LLM)-assisted extraction was applied to identify PK parameters from unstructured DrugBank text descriptions. Bioavailability (F) values were extracted using rule-based methods for 79 compounds and supplemented by LLM-assisted extraction for an additional 60 compounds. If oral bioavailability data are unavailable, bioavailability is assumed to be 1. Likewise, the missing fu is assumed to be 1 as well. Absorption rate constants (*k_a_*) were frequently unavailable and were therefore calculated from reported time to maximum concentration (*t_max_*) values, assuming first-order absorption and disposition (Equation 1):

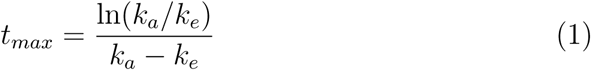

where *k_e_* is the elimination rate constant, which can be calculated by *CL/V_d_*.

Approved drug entries were obtained from ChEMBL by selecting parent compounds that had reached Phase 4 (i.e., marketed drugs), along with their chemical structures and physicochemical properties. Drug physicochemical properties were used to calculate tissue partition coefficients for lung, liver, and kidney using published methods [29, 30].

Virus information in PPP-db was extracted from Ianevski *et al.* [26], which includes virus families, virus groups, and virus names. Antiviral bioassay data were extracted from ChEMBL by linking approved drugs with virus entries including *IC*_50_ and *EC*_50_ values.

## 3. PPP-COMPUTE workflow

PPP-COMPUTE provides three corresponding user workflows, PK, PD, and clinical trial design evaluation, that guide users step by step through each module (Figure 3) building on the three core modules described above. Supported by PPP-db, these workflows are described and demonstrated using a representative example in the following subsections.

**Figure 3:**
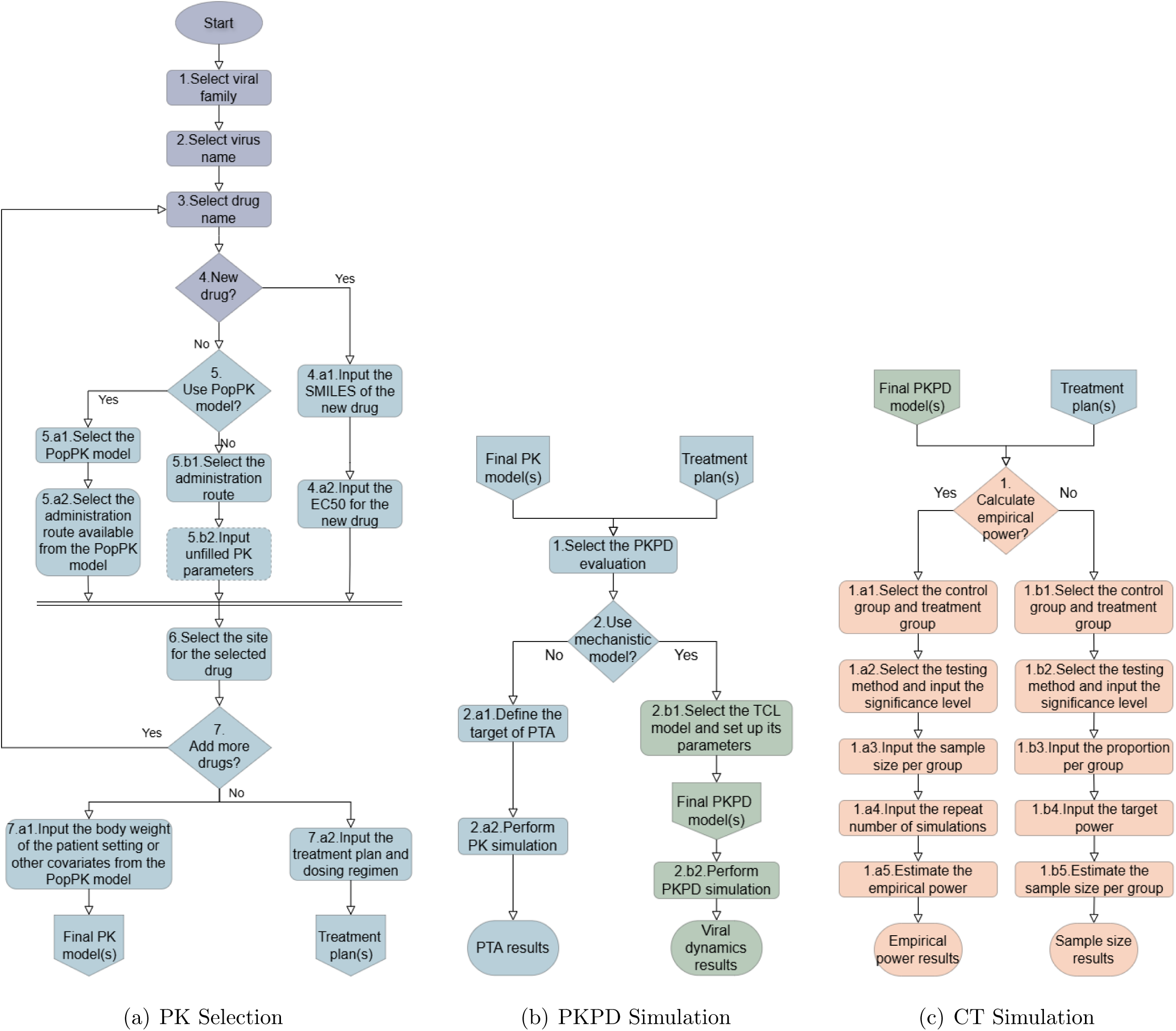
Detailed workflow of PPP-COMPUTE. The oval indicates the starting point of the workflow, and circle denotes connector to subsequent workflows. Diamond represents user decision points, and rounded rectangle indicates actions.

### 3.1. PK morkflom

Users begin PPP-COMPUTE with the PK workflow. The workflow is illustrated in Figure 3a with numbered steps.

#### 3.1.1. Virus and drug selection

The user selects a virus family and virus name (Steps 1–2). Based on the selected virus, the platform retrieves from PPP-db the list of approved drugs that have been tested against that virus (Step 3). If the user selects “New Virus”, the available drug list is expanded to include drugs tested against viruses from the same viral family, and their bioassay records are pooled to provide antiviral potency values for subsequent simulations (Figure 4).

**Table 1:** The statistics of the PPP-db.

| Item | Description | No. Entries |
| --- | --- | --- |
| Drug substances | Approved drugs extracted from ChEMBL | 2384 |
| Virus family | Virus families classified based on genetic and biological characteristics | 19 |
| Virus | Virus names included in the database | 54 |
| $IC_{50}/EC_{50}$ | Drug concentrations required to achieve 50% inhibition or antiviral effect against a given virus | 103193 |
| PK parameters | PK parameters extracted from DrugBank | 1424 |
| Oral PK parameters | First-order oral absorption parameters extracted from DrugBank | 258 |
| Predicted PK parameters | PK parameters predicted using QSPR models | 2228 |
| Target partition | Partition coefficients from free plasma to target tissues | 715 |
| Dosing regimen | Approved dosing regimens extracted from drug labels and reference sources | 573 |
| PopPK model library | Population PK models implemented from published studies | 19 |

**Figure 4:**
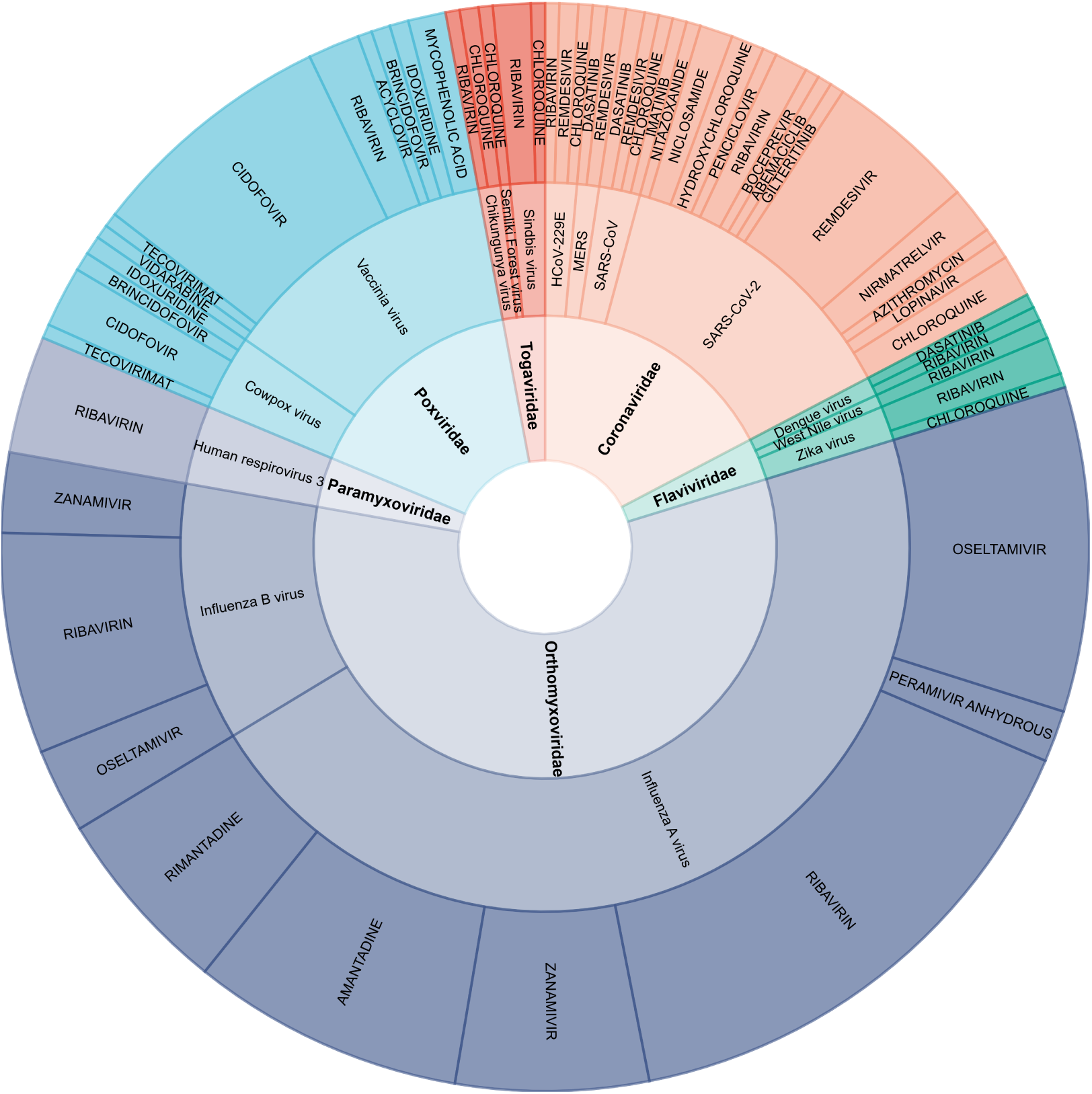
Overview of virus-drug combinations in PPP-db for the top six viral families with pandemic potential included in the database. The chart has three concentric rings. The inner ring represents viral families, color-coded by family. The middle ring shows individual viruses within each family. The outer ring displays approved drugs with documented antiviral activity against the corresponding virus. Only virus-drug combinations supported by more than three bioassay records from ChEMBL (Version 34) [18] are shown. Segment width is proportional to the number of bioassay records for that virus-drug pair.

*Example*: As an example, Severe Acute Respiratory Syndrome Coronavirus 2 (SARS-CoV-2) is selected, which retrieves the corresponding antiviral drug list from PPP-db.

#### 3.1.2. PK prediction for nem drug candidates

The “Drug Selection” option allows users to evaluate either a new compound or an approved drug (Step 4). For a “New Drug”, the user provides the SMILES string and the corresponding *EC*_50_ value (Steps 4.a1–4.a2). Pharmacokinetic parameters including volume of distribution (Vd), clearance (CL), and free fraction (fu) are predicted from the SMILES structure using previously developed QSPR models [16, 17]. For new compounds, the administration route is limited to intravenous dosing and drug concentrations are simulated in plasma.

*Example*: As an example, remdesivir is evaluated as a new drug candidate against SARS-CoV-2. The SMILES string, *in vitro EC*_50_ value, and dosing regimen are listed in Table 2. PK parameters (Vd, CL, and fu) are predicted using the QSPR model, and the resulting simulated free plasma concentrations are shown in Figure 5.

**Figure 5:**
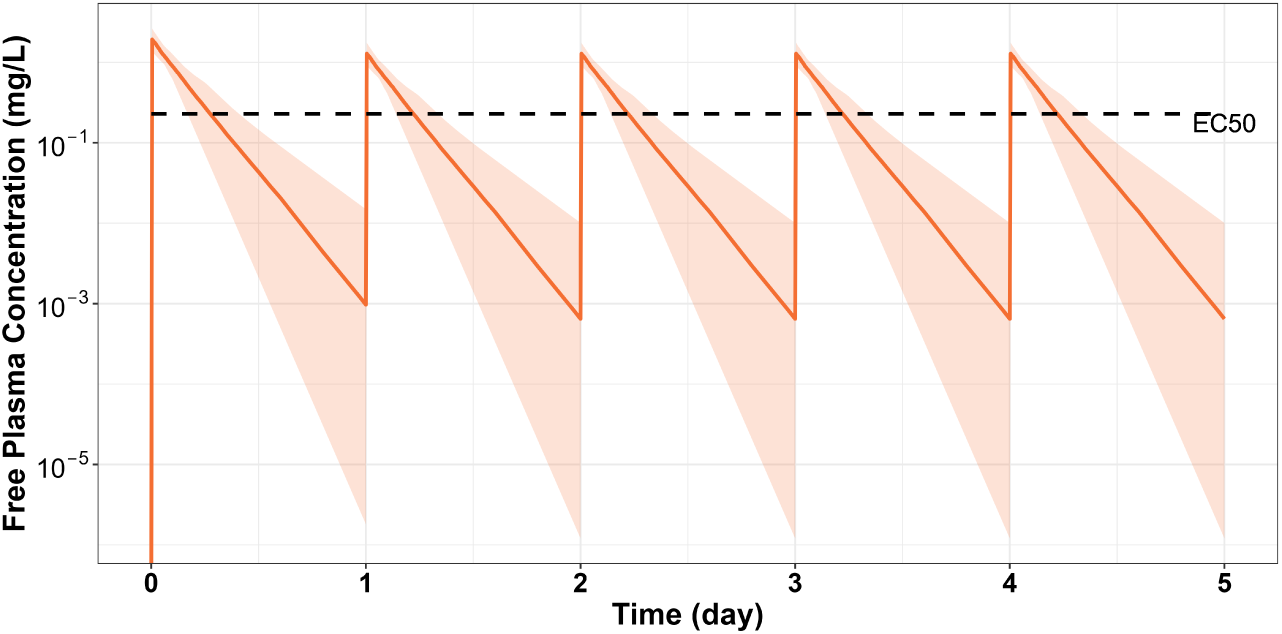
Free drug concentration plasma profile for a “New Drug” with PK parameters predicted by the QSPR model (volume of distribution, clearance, and protein binding). The dosing regimen follows Table 2. The solid orange line represents the median drug concentration, and the shaded area indicates the 95% prediction interval from simulations of 100 individuals, with IIV in clearance and volume of distribution of *ω*^2^ = 0.04. The dashed line indicates the median *in vitro EC*_50_ listed in Table 2.

**Table 2:** Drug information (remdesivir)

|  |  |
| --- | --- |
| Drug Name | Remdesivir |
| SMILES | CCC(CC)COC(=O)[C@H](C)N[P@](=O)(OC[C@@H]1[C@H]([C@H]([C@](O1)(C#N)C2=CC=C3N2N=CN=C3N)O)O)OC4=CC=CC=C4 |
| $EC_{50}$ (mg/L) | 0.2290 |
| Dosing Regimen | Once per day, 3 mg/kg on Day 1 and 2 mg/kg on Day 2 to 5. |

#### 3.1.3. Approved drug PK simulation

For approved drugs, PK simulations can be performed using either NCA-derived PK parameters or published population PK models, both stored in PPP-db (Step 5). If NCA-derived PK parameters are used (Steps 5.b1–5.b2), the administration route can be specified as intravenous or oral, with intra-venous dosing as the default. Drug disposition is described using a one-compartment model with linear clearance, and oral dosing additionally assumes linear absorption. Intravenous simulations require Vd, CL, and fu, while oral simulations additionally require bioavailability (F) and the absorption rate constant (ka). PK parameters derived from *in vivo* studies are available and antiviral potency values can be retrieved directly from PPP-db. Simulated concentration–time profiles can therefore be evaluated relative to the corresponding *EC*_50_ values.

*Example*: Using remdesivir as an example, PK simulations based on NCA-derived PK parameters produce the concentration–time profile shown in Figure 6A.

**Figure 6:**
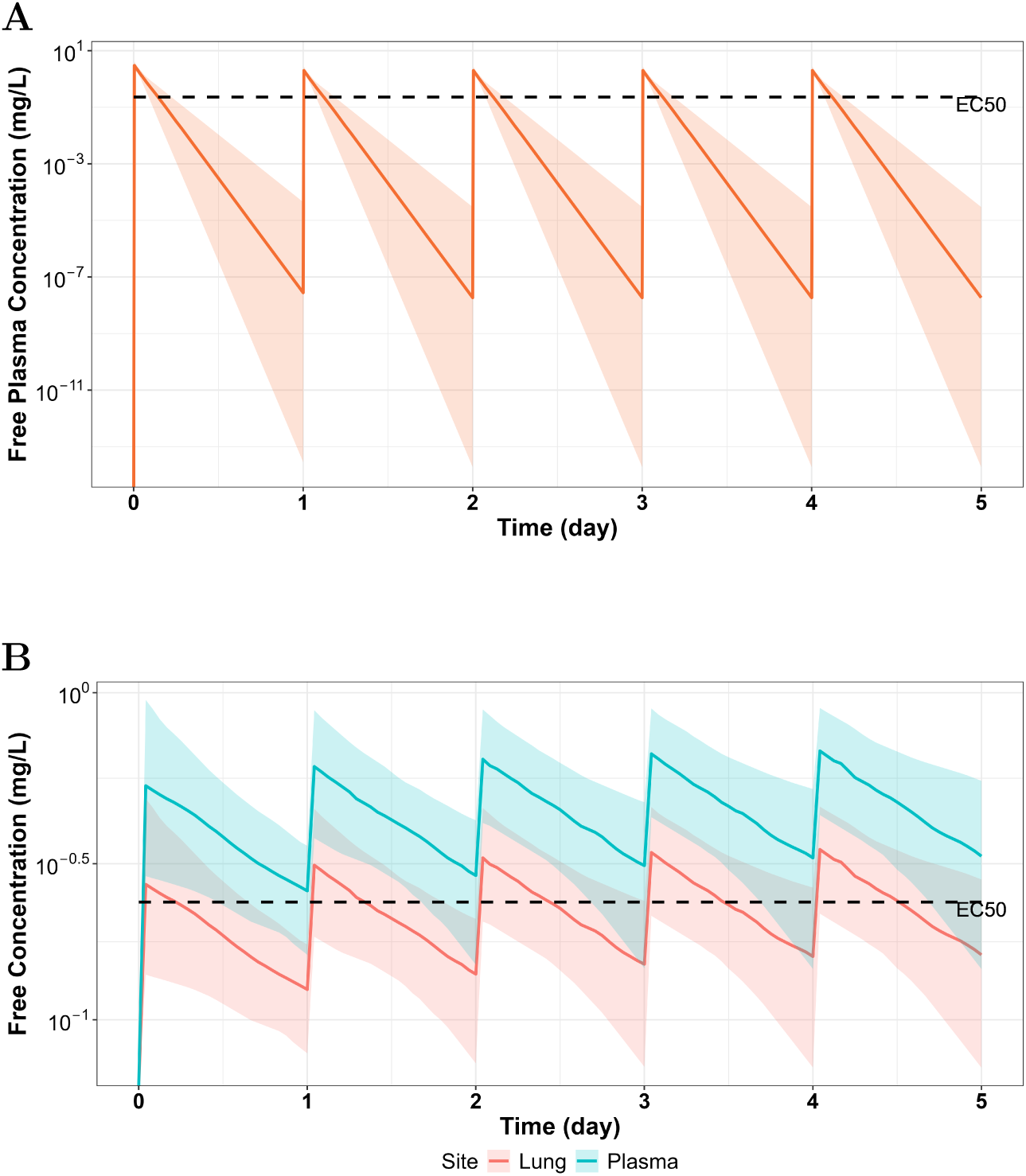
Free plasma concentration profiles of remdesivir following the dosing regimen in Table 2, simulated using PK parameters retrieved from PPP-db (A) or the published PopPK model from Sukeishi et al. [32] (B). Solid lines represent median predicted concentrations and shaded areas indicate 95% prediction intervals based on 100 simulated individuals. In panel A, inter-individual variability in clearance and volume of distribution is characterized by a variance of *ω*^2^ = 0.04. In panel B, inter-individual variability follows the published model, and orange and blue curves represent free drug concentrations in the lung and plasma, respectively. The dashed line denotes the median *in vitro EC*_50_.

Alternatively, users may select a PopPK model from the library (Steps 5.a1–5.a2), if available. The PopPK model library includes published model structures, parameter values, covariate relationships, and population characteristics of the source population for each available drug. Any required covariates are automatically added to the Patient Settings interface.

#### 3.1.4. Final PK model and treatment plan

Users may simulate drug concentrations at target sites including lung, liver, and kidney (Step 6). Tissue concentrations are estimated by scaling plasma concentrations using tissue partition coefficients available in PPP-db. At this stage, the base PK model is defined, consisting of either a one-compartment model with NCA-derived or QSPR-predicted parameters, or a literature PopPK model.

In Step 7, users define the final PK models and treatment plans. Up to five drugs can be simulated simultaneously. Population characteristics (shared for all treatment plans) can be adjusted via model covariates to reflect the target population (Step 7.a1). Dosing regimens extracted from PPP-db can be selected or modified for each drug (Step 7.a2). These treatment plans are passed to the PD and CT workflows as well for further evaluation.

*Example*: Using the PopPK model for remdesivir selected from the PPP-db model library [32], the lung is specified as the target site of interest. The source population of this model has a mean age of 72 years and an eGFR of 74.7 ml/min. Using the treatment plan in Table 2, the population characteristics are adjusted to reflect a younger population with a mean age of 60 years and an eGFR of 90 ml/min. The resulting plasma and lung concentration-time profiles are shown in Figure 6.

### 3.2. PD morkflom

The defined final PK models and treatment plans are in the next step used in the PD workflow to evaluate antiviral activity through PTA analysis or simulation of viral dynamics (Step 1 in Figure 3b).

#### 3.2.1. Probability of Target Attainment Analysis

For PTA analysis, users specify the fraction of treatment time during which the free drug concentration exceeds the target potency value (% fT above *EC*_50_) and select the target *EC*_50_ value (Step 2.a1). When “New Virus” is selected, the default *EC*_50_ corresponds to the median value from pooled bioassay records for viruses within the relevant family, which for this example is the *Coronaviridae* family. Alternatively, users may specify a userdefined *EC*_50_. Based on the selected treatment plan, drug concentrations are simulated and used to calculate PTA values (Step 2.a2).

*Example*: Using the remdesivir PopPK model defined in the PK workflow, PTA results are shown in Figure 7. The target potency can be specified as either *EC*_50_ or *EC*_90_, and PTA curves are generated across a range of these values. The PTA percentage represents the proportion of individuals for whom at least 50% of the treatment time the free drug concentration exceeds the target *EC*_50_. As shown in Figure 7, approximately 90% of individuals achieve 50% fT above the *EC*_50_ in plasma, while lower attainment is observed for lung concentrations. These results suggest that remdesivir may achieve effective exposure for a substantial proportion of patients.

**Figure 7:**
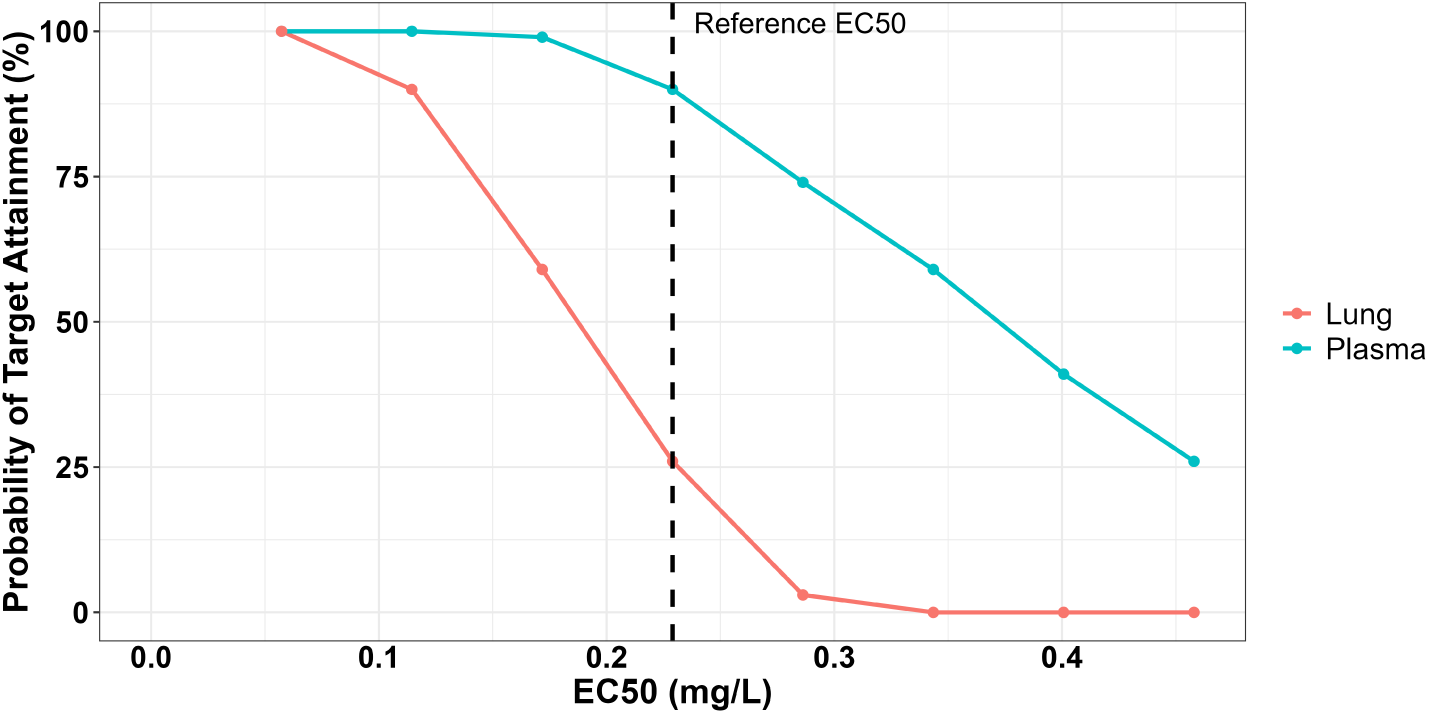
PTA (50% fT *≥ EC*_50_) curves for remdesivir in both plasma and lung based on PK simulation shown in Figure 6B. PTA is defined as the proportion of individuals for whom the free drug concentration exceeds the target *EC*_50_ for at least 50% of the treatment period.

#### 3.2.2. Viral Dynamic Simulations

Antiviral effects can also be evaluated using simulations of viral dynamics using the mechanistic viral dynamic model (Step 2.b1). Users can specify the initial conditions of the model, including the initial number of target cells (*T*_0_) and the initial viral load (*V*_0_), thereby defining the final PK/PD model. Simulations are then performed using the selected treatment plans (Step 2.b2). The resulting viral dynamic curves allow users to evaluate viral replication and treatment effects simultaneously and to derive virological endpoints, including viral load at the end of treatment and the area under the viral dynamic curve (AUC).

*Example*: Viral dynamic simulations for remdesivir under different *EC*_50_ values and treatment delays are shown in Figure 8. The PK input corresponds to the free plasma concentrations simulated in Figure 6B. The viral dynamic curves illustrate how antiviral treatment influences viral clearance.

**Figure 8:**
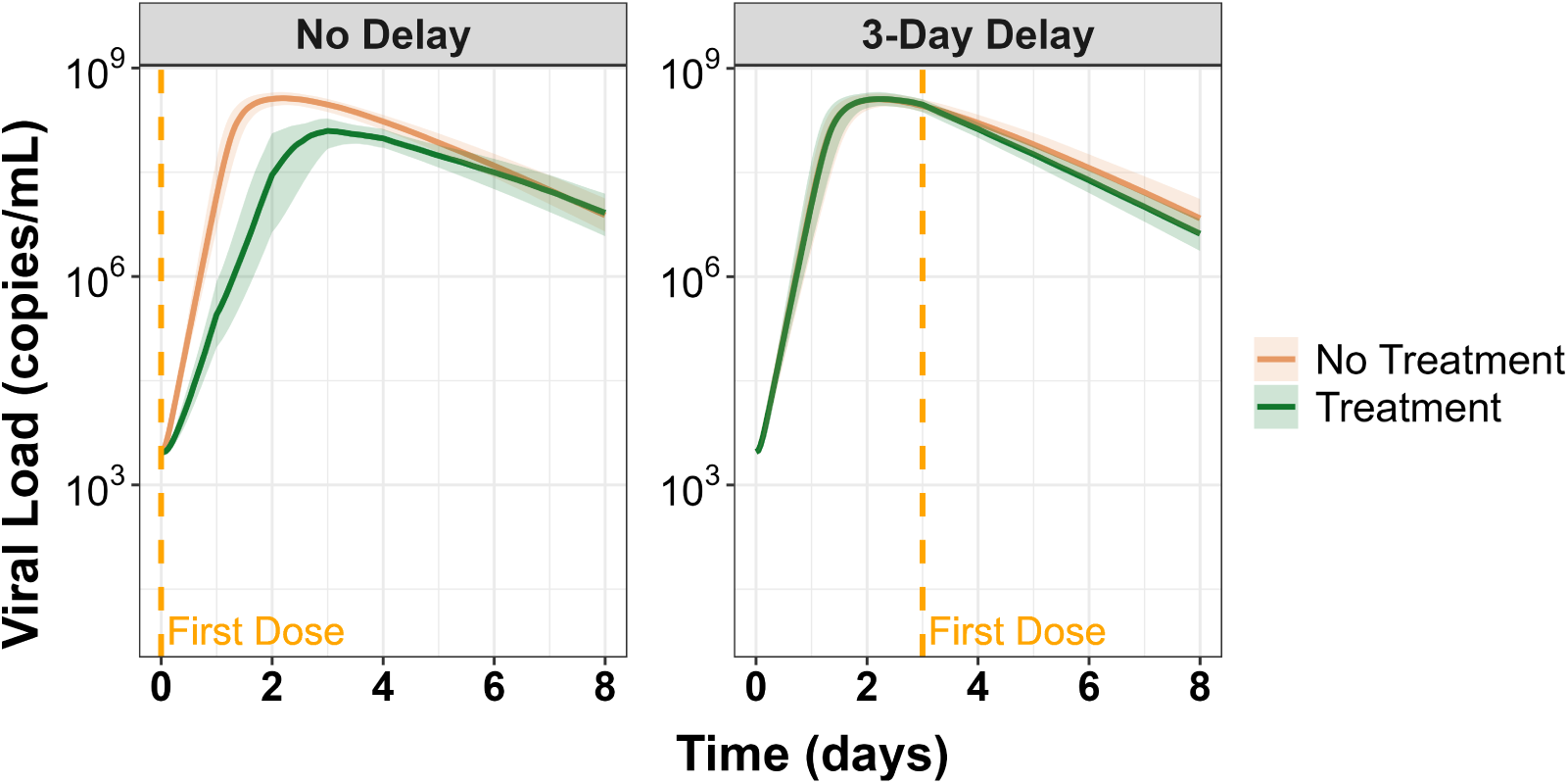
Viral dynamic simulations for remdesivir under different *EC*_50_ and treatment initiation time (no delay or 3-day delay). PK input is based on the free plasma drug concentration shown in Figure 6B. IIV for infection rate *β*, cell death rate *δ*, and virus production rate *ρ* is set to *ω*^2^ = 0.01. Orange curves represent simulations without treatment, whereas green curves represent treatment with remdesivir. The vertical dashed line indicates the start of treatment.

### 3.3. Clinical trial design evaluation morkflom

After completing the PD workflow, users can proceed to clinical trial design as illustrated in Figure 3c. Based on the final PK/PD models and treatment plans, the CT workflow supports sample size estimation and empirical power analysis. By integrating PK simulations, viral dynamic modeling, and statistical evaluation, PPP-COMPUTE enables quantitative evaluation of candidate drugs from predicted exposure and antiviral response to expected clinical trial performance.

#### 3.3.1. Sample size estimation

For sample size estimation (Steps 1.b1–1.b4), users assign treatment plans to control and treatment groups and specify the allocation ratio between groups. The default control group is defined as “No Treatment”. Users then select the statistical test, significance level (default 5%), and target power (default 80%). Sample sizes are estimated by simulating virological endpoints and performing statistical testing with Bonferroni correction [33].

*Example*: The clinical trial design workflow is demonstrated using remdesivir with the same PK/PD model defined previously. The control group is specified as “No Treatment” and the treatment group as “Treatment”, corresponding to remdesivir with the dosing regimen defined in Table 2. The associated viral dynamic curves are shown in Figure 8. Sample size can be estimated from the log_10_-transformed area under the viral dynamic curve or viral load at the end of treatment. Monte Carlo simulations are used to generate virological endpoints for a target power of 80%. In the current example, the estimated sample size of 3-day delayed treatment is four subjects per group using the viral load at the end of treatment with equal allocation between control and treatment groups.

#### 3.3.2. Empirical pomer analysis

Users can perform empirical power analysis (Steps 1.a1–1.a4) by specifying the treatment plans for the control and treatment groups together with the statistical test and significance level. Instead of defining an allocation ratio, users provide the cohort size for each group, and empirical power is calculated based on PK/PD simulations.

*Example*: Using the same remdesivir example, empirical power analysis is performed with the predicted viral dynamic curves shown in Figure 8 with 3-day delayed treatment. When the virological endpoint is defined as the log_10_-transformed viral load at the end of treatment, the empirical power reaches 80% at a sample size of 4, consistent with the target power used in the sample size estimation. The effect size calculated using Cohen’s *d* is *−*1.3408 with a 95% confidence interval of (*−*3.2556, 0.5740). Statistical testing is performed using Welch’s *t*-test, which assumes normally distributed endpoints, although the Mann–Whitney U test is also available when distributional assumptions are not desired.

## 4. Discussion

PPP-COMPUTE is designed to support rapid evaluation of repurposed drugs and novel compounds and to inform clinical trial design during an epidemic or a pandemic within an accessible web-based platform. The integration of PK, PD, and clinical trial design tools allows users to assess whether candidate drugs with potential antiviral activity *in vitro* are likely to achieve effective exposure *in vivo*. In addition, the platform helps translate these predictions into quantitative insight and further into treatment benefit and clinical study design. This platform supports long-term preparedness across a broad range of potential pandemic pathogens as multiple virus families were included.

PPP-COMPUTE complements existing drug-repurposing platforms as it addresses practical questions relevant to a pandemic response. Many current tools prioritize candidate drugs based on molecular interactions or network associations, whereas PPP-COMPUTE evaluates whether candidate drugs can plausibly achieve effective exposure and antiviral responses under clinically realistic conditions. The platform guides how antiviral activity may translate into virological endpoints across different dosing regimens and treatment scenarios by integrating antiviral potency data with mechanistic PK/PD simulations. The clinical trial design module builds on these simulations by linking predicted antiviral responses to statistical analyses. This enables the calculation of sample size and empirical power under different study assumptions and supports translation into clinical trial design. This unified workflow supports evaluation from early candidate selection and dosing feasibility to clinical trial design. This is all achieved within a single platform, which provides practical decision support for researchers, clinicians, and trial designers involved in pandemic preparedness.

Virological endpoints provide early quantitative evidence of antiviral activity and support rapid evaluation of candidate therapies during outbreak situations. Viral load dynamics, viral clearance rates, and viral load area under the curve are indicators of treatment efficacy before clinical outcomes are observable. Studies have shown that viral dynamics are associated with clinical progression, as faster viral clearance has been linked to improved outcomes [34, 35], and viral load has been shown to correlate with antiviral exposure and treatment response [36]. These findings support the use of virological endpoints as surrogate measures for early drug evaluation and dose selection. The relationship between viral dynamics and clinical outcomes is highly variable between different pathogens and disease settings, thus virological endpoints should be interpreted as indicators of antiviral activity rather than direct predictors of clinical benefit.

PPP-db provides the data foundation for the platform by linking approved drugs with virus information, PK parameters, dosing regimens, and antiviral bioassay data. PPP-db includes inputs required for PK/PD simulations and reduces the manual effort needed to assemble datasets during outbreak situations by harmonizing data from multiple open sources. In contrast to many existing databases [18, 19, 20, 21, 22, 23, 24, 25, 26, 27, 28], PPP-db is specifically structured to support PK/PD simulations by integrating PK parameters, dosing regimens, PopPK models, mechanistic PK/PD models, and antiviral potency data within a single framework. These types of data are often scattered across multiple resources and publications, and their integration within PPP-db facilitates consistent and reproducible PK/PD simulations. The database includes a broad range of FDA-approved drugs beyond antivirals, which allows PPP-db to be used as a standalone resource for pharmacological and translational research. The FAIR-oriented structure of PPP-db supports reproducibility and future expansion as new data become available.

PPP-COMPUTE is publicly accessible and developed within an open-science framework to support transparent, reproducible, and collaborative research. The web-based interface enables users without specialized modeling expertise to perform PK/PD simulations and clinical trial evaluations, which lowers the barrier to quantitative pharmacological analysis during outbreak situations. Open access to the platform and PPP-db facilitates continuous improvement through community contributions, allowing models and datasets to accumulate and evolve as new experimental and clinical evidence is available.

PPP-COMPUTE has several limitations that should be considered when using the platform and interpreting its results. PK simulations based on NCA-derived or QSPR-predicted parameters rely on simplified assumptions, including representation of drug disposition with a one-compartment model and linear elimination. When oral bioavailability data are unavailable, bioavailability is assumed to be 1, which may overestimate systemic exposure for some compounds. For new compounds, PK parameters are predicted using QSPR models to support early feasibility assessment when experimental data are unavailable. However, accuracy depends on the quality and variability of the training data and may be reduced for compounds outside the model’s applicability domain. PPP-db integrates pharmacological and virological information from multiple public sources, introducing heterogeneity in reported PK parameters and antiviral potency values. Systematic curation and automated extraction have improved consistency, however variability of different sources may still affect the reliability of parameters. A further limitation concerns IIV. The simulations use relatively low IIVs when no literature model is available (e.g., the example simulations), which may inflate statistical power. In such cases, the resulting sample size and power estimates should be viewed as optimistic lower bounds. Users are advised to specify realistic IIV to obtain more conservative estimates. Despite these limitations, PPP-COMPUTE enables rapid PK/PD simulations with limited available data, which are particularly valuable in early outbreak settings where timely decisions must be made.

PPP-COMPUTE is a flexible platform that can evolve with advances in PK/PD modeling, data availability, and outbreak preparedness strategies. Future developments may include expansion of virus-specific PK/PD model libraries and incorporation of disease-specific data to strengthen the link between viral dynamics and clinical outcomes. The platform currently focuses on viral pathogens, however its modular structure makes extension to other infectious organisms, such as bacterial, fungal, or parasitic pathogens, relatively straightforward. The pathogen-independent PK module carries over directly, while the potency measures, dynamic models, and endpoints can be replaced by organism-specific equivalents, such as the minimum inhibitory concentration (MIC) and bacterial growth and kill models. The framework could also be extended to other therapeutic modalities. For instance, it could in principle inform the design of vaccine trials by coupling the simulation engine to models of host immune response. Together, these directions would broaden PPP-COMPUTE from an anti-infective treatment tool to a more general platform for model-informed evaluation of pandemic interventions.

## 5. Conclusion

We developed PPP-COMPUTE, a publicly accessible PK/PD simulation platform, to support rapid evaluation of repurposed and novel anti-infective candidates and clinical trial design during emerging pandemics. The platform provides a structured environment that supports researchers and clinicians in pandemic preparedness.

## Data Availability

All data produced are available online at https://github.com/LeidenPharmacology/pppcompute

## Code availability

The source code of PPP-COMPUTE is publicly available at https://github.com/LeidenPharmacology/pppcompute. The platform is accessible at https://pppcompute.lacdr.leidenuniv.nl/.

