## Supplementary Material for "PPP-COMPUTE: A Pandemic Pharmacology Platform for COMPUTational Evaluation of Anti-infectives in Pandemic Preparedness"

**Tingjie Guo^*^**

*Leiden Academic Centre for Drug Research (LACDR), Leiden University, Einsteinweg 55, Leiden, 2333 CC, The Netherlands*

**^*^Contributed equally and share senior authorship**

**Correspondence:**

Coen van Hasselt

Linda Aulin

Tingjie Guo

### Section S1. PPP-COMPUTE Platform screenshot

In this section, the PPP-COMPUTE platform UI will be provided, including the detailed sidebar and main page sections. The PPP-COMPUTE UI is shown in Figure S1. The summarized results or clinical trial design results are shown on the default page of PPP-COMPUTE, as shown in Figure S1a. The separated treatment pages include the information on the PK curves and the corresponding PTA or viral dynamics curves, shown in Figures S1b and S1c.

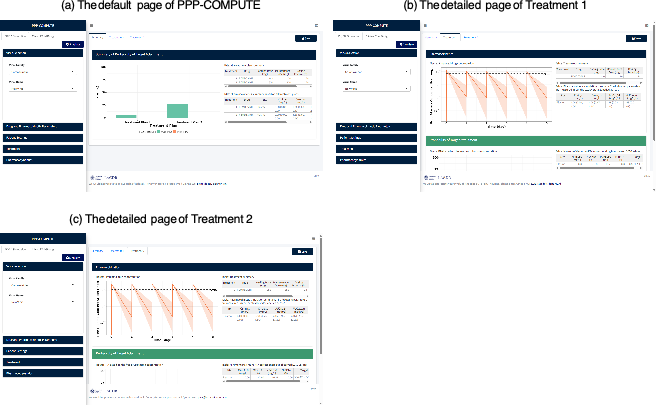

**Figure S1:** The overall platform UI screenshot of PPP-COMPUTE. The screenshot is from an example of discovering 2 dosing regimens (Treatments) using remdesivir against a new virus in the family Coronaviridae.

The sidebar of PPP-COMPUTE consists of “PK/PD Simulation” and “Clinical Trials” tabs. The “PK/PD Simulation” includes 5 sections: Virus Selection, Drug and Pharmacokinetic Parameters, Patient Settings, Treatment, and Pharmacodynamics, and the “Clinical Trial Design” includes 2 sections: Sample Size Estimation and Power Analysis. The details of each section are shown in Figure S2. In the example in Figure S2, it showcases the sidebar with drug NCA PK parameters.

If the user selects Model Library (shown in Figure S2b), the Drug and Pharmacokinetic Parameters and Patient Settings sections are changed and shown in Figure S3. The PK parameters follow the selected PK model from the literature, resulting in added covariates under the Patient Settings, shown in Figure S3b.

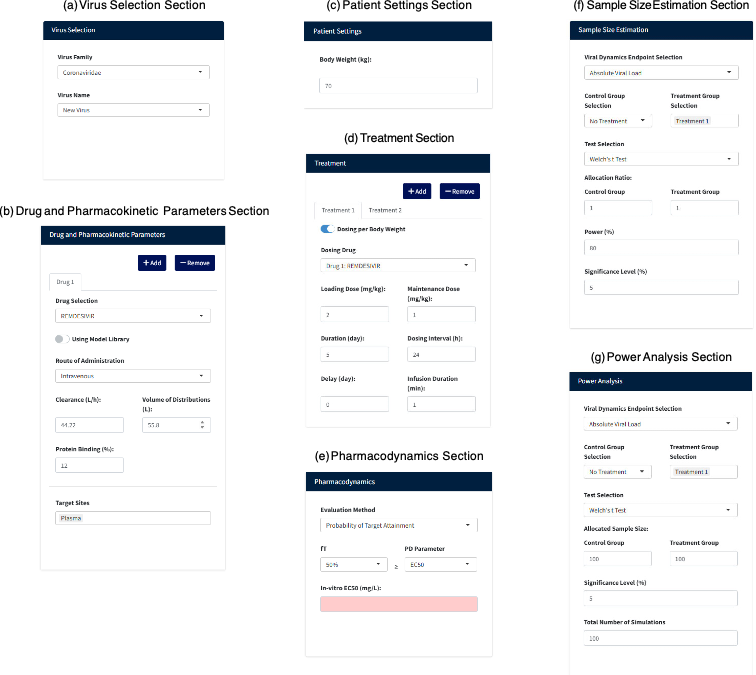

**Figure S2:** The detailed sidebar screenshot of PPP-COMPUTE. Panels (a) – (e) show the sections for PK/PD Simulation, while Panels (f) and (g) show the sections for Clinical Trial Design.

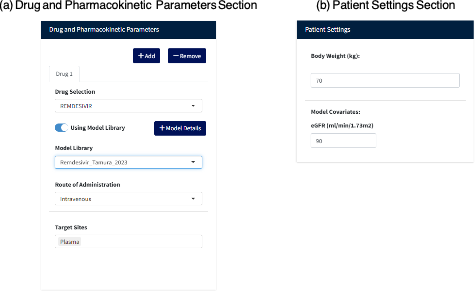

**Figure S3:** The updated sections of Drug and Pharmacokinetic Parameters and Patient Settings when selecting Model Library.

### Section S2. Details of PPP-db

PPP-db is curated from multiple sources, including ChEMBL (Version 34), DrugBank (Version 5.1.12), DailyMed, openFDA, Drugs.com, Medscape, and Farmacotherapeutisch Kompas, as well as published studies, namely Ianevski et al. (2022), Lombardo et al. (2018), and Mamada et al. (2021). This PPP-db is used to support the PPP-COMPUTE workflows. In this section, the type of each column is labeled in CHAR (character), INT (integer), FLOAT (float), and BOOL (boolean). The key column is listed after K, and the foreign key is listed after FK. The ER diagram of PPP-db is shown in Figure S4.

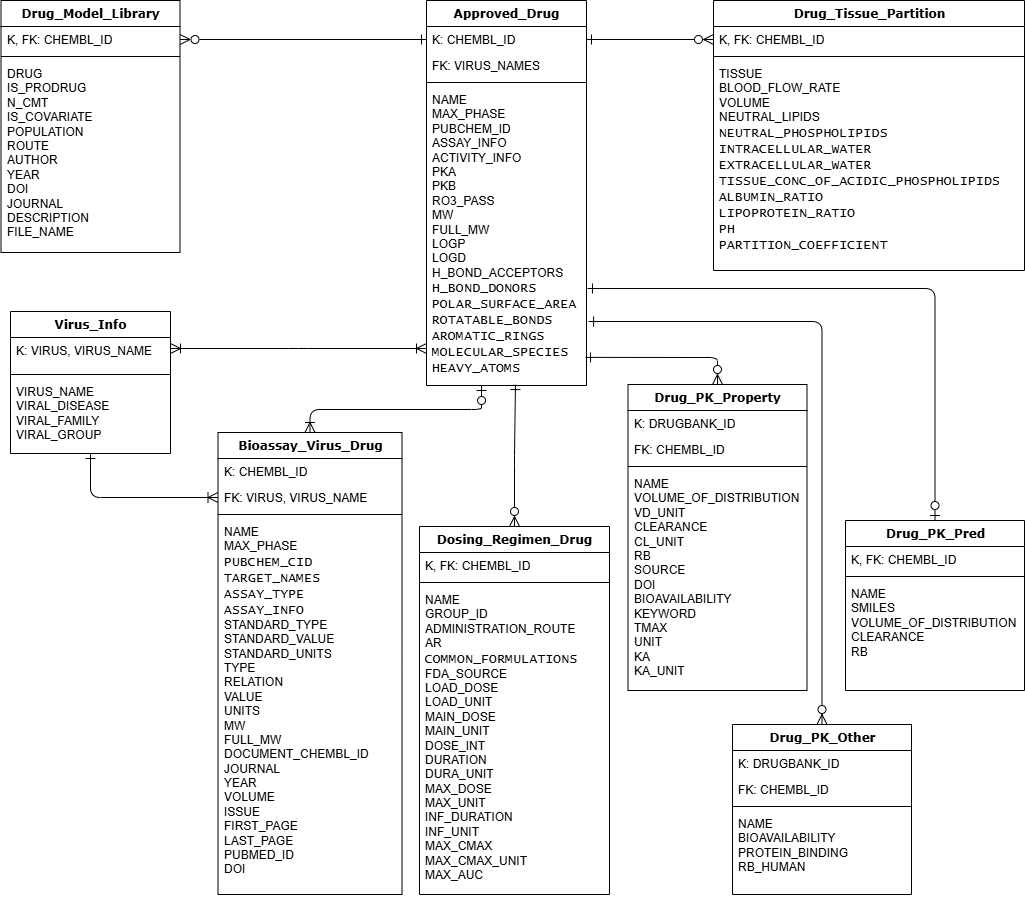

**Figure S4:** The comprehensive schema of the PPP-db. The tables contained in PPP-db are shown in the squares, with the key column listed after K, and the foreign key listed after FK. The relation with a dash on one side means “One” entity on this side. The circle represents the “Zero” entity in the table. The fork symbol means “Many” entities on this side. The combinations of the aforementioned symbols, as dash-circle, circle-fork, and dash-fork, represent “zero-or-one”, “zero-or-many”, and “one-or-many”, respectively.

### S2.1 Virus Information

The virus information is contained in the “Virus_Info” table. The data are extracted from DrugVirus.info 2.0. We extracted the virus name, viral disease, viral family, and viral group from the BSA table by uniquely identifying each virus. Therefore, we can identify the unique viruses and the diseases they cause, along with their families and groups.

From the BSA table of DrugVirus.info 2.0, we obtained the summary statistics listed in Table 1. Both the virus and the virus name are used to retrieve records from the “Bioassay_Virus_Drug” table.

Table 1: The summary statistics of the Virus_Info

| Column | Type | description | Number of unique records |
| --- | --- | --- | --- |
| Virus (K) | CHAR | The abbreviated form of virus name | 64 |
| Virus name (K) | CHAR | The full name of virus | 54 |
| Viral disease | CHAR | The disease caused by the virus | 51 |
| Viral family | CHAR | A group of viruses based on a combination of genetic and biological properties of the viruses | 19 |
| Viral group | CHAR | The Baltimore classification system for the virus | 6 |

Table 2: The SQL for extracting the approved drugs from ChEMBL

SELECT DISTINCT

parent.chembl_id AS compound_id,

parent.pref_name AS Name,

md.max_phase,

cp.cx_most_apka AS pKa,

cp.cx_most_bpka AS pKb,

cp.cx_logp AS logP,

cp.cx_logd AS logD,

cp.RO3_PASS AS RO3_PASS,

cp.MW_FREEBASE AS MW,

cp.full_mwt AS full_MW,

cp.hba AS H_bond_acceptors,

cp.hbd AS H_bond_donors,

cp.psa AS polar_surface_area,

cp.rtb AS rotatable_bonds,

cp.aromatic_rings,

cp.molecular_species,

cp.heavy_atoms

FROM molecule_dictionary md

JOIN molecule_hierarchy mh ON md.molregno = mh.molregno

JOIN molecule_dictionary parent ON mh.parent_molregno = parent.molregno

JOIN activities a ON md.molregno = a.molregno

JOIN assays ass ON a.assay_id = ass.assay_id

JOIN target_dictionary t ON ass.tid = t.tid

LEFT JOIN activity_properties ap ON a.activity_id = ap.activity_id

LEFT JOIN compound_properties cp ON parent.molregno = cp.molregno

LEFT JOIN compound_records cr ON parent.molregno = cr.molregno

LEFT JOIN source src ON cr.src_id = src.src_id

LEFT JOIN compound_structures cs ON parent.molregno = cs.molregno

WHERE md.max_phase = 4

GROUP BY parent.molregno

ORDER BY md.max_phase DESC, parent.pref_name;

### S2.2 Approved Drug

Information on approved drugs is retrieved from ChEMBL. The SQL to extract approved drugs and their compound properties is listed in Table 2. In this table, we extracted the physiological and structural properties. The detail of the “Approved_Drug” table is listed in Table 3.

Table 3: The summary statistics of the Approved_Drug

| Column | Type | Description | Number of unique records |
| --- | --- | --- | --- |
| chembl_id (K) | CHAR | The unique identifier defined by ChEMBL for a drug | 2384 |
| Name | CHAR | The name of a drug | 2384 |
| max_phase | INT | The phase of the drug development. ChEMBL defines 4 for an approved drug. | 2384 |
| PubChem_CID | CHAR | The unique identifier defined by PubChem for a drug | 2367 |
| Virus_Names (FK) | CHAR | The group of viruses treated by a drug | 2384 |
| assay_info | CHAR | The group of assays used by a drug | 2384 |
| activity_info | CHAR | The group of bioactivity measurements used by a drug | 2315 |
| pKa | FLOAT | The negative logarithm of the acid dissociation constant | 1306 |
| pKb | FLOAT | The negative logarithm of the base dissociation constant | 1280 |
| RO3_PASS | CHAR | The Rule of 3 pass to identify fragment-like compounds that are more likely to be hits in fragment screening | 2146 |
| MW | FLOAT | The parent form of the molecule after removing any salts | 2325 |
| full_MW | FLOAT | The molecular weight of the full compound, including any salts or hydrates present | 2325 |
| logP | FLOAT | The measure of the lipophilicity of a neutral molecule | 2146 |
| logD | FLOAT | The measure of lipophilicity, including ionized species at a given pH | 2146 |
| H_bond_acceptors | INT | Hydrogen bond acceptors | 2146 |
| H_bond_donors | INT | Hydrogen bond donors | 2146 |
| polar_surface_area | FLOAT | The combined surface area of all polar atoms in the molecule | 2146 |
| rotatable_bonds | INT | The number of single non-ring bonds attached to non-terminal heavy atoms that can rotate freely | 2146 |
| aromatic_rings | INT | The count of ring systems in the molecule that are aromatic | 2146 |
| molecular_species | CHAR | The chemical form of the drug molecule | 2104 |
| heavy_atoms | INT | The count of all atoms in the molecule except hydrogens | 2146 |

### S2.3 Bioassay Virus Drug

Information on approved drugs is retrieved from ChEMBL. The SQL to extract approved drugs and their compound properties is listed in Table 4. From this SQL, we can extract the IC50/EC50 and Emax from the in-vitro experiments (stated as the “F” assay type, which represents functional assays). As stated in the ChEMBL documents (https://chembl.gitbook.io/chembl-interface-documentation/frequently-asked-questions/chembl-data-questions), Type “B” (binding studies) can also contain IC50 information. Therefore, the SQL in Table 4 can also be run without the assay-type condition.

To comprehensively capture the antiviral drugs bioassays, we have also applied the SQL from Table 5 with the drug chembl_id. We can get the list of drugs that are not covered from the SQL in Table 4. We can loop over the chembl_ids in this list and extract the IC50/EC50 values using the SQL in Table 5.

Once we obtained the joined tables from the SQL queries in Tables 4 and 5, we filtered out records for viruses not in the virus list.

Therefore, we constructed the bioassay table by combining virus and drug information. The summary statistics of the “Bioassay_Virus_Drug” are listed in Table 6.

Table 4: The SQL for extracting the bioassays from ChEMBL

SELECT DISTINCT

parent.chembl_id AS chembl_id,

parent.pref_name AS Name,

md.max_phase,

cr.compound_key AS PubChem_CID,

t.pref_name AS target_names,

ass.assay_type,

ass.description AS assay_info,

a.standard_type,

a.standard_value,

a.standard_units,

ap.type,

ap.relation,

ap.value,

ap.units,

cp.MW_FREEBASE AS MW,

cp.full_mwt AS full_MW,

docs.chembl_id AS document_chembl_id,

docs.journal AS journal,

docs.year AS year,

docs.volume AS volume,

docs.issue AS issue,

docs.first_page AS first_page,

docs.last_page AS last_page,

docs.pubmed_id AS pubmed_id,

docs.doi AS doi

FROM molecule_dictionary md

JOIN molecule_hierarchy mh ON md.molregno = mh.molregno

JOIN molecule_dictionary parent ON mh.parent_molregno = parent.molregno

JOIN activities a ON md.molregno = a.molregno

JOIN assays ass ON a.assay_id = ass.assay_id

JOIN target_dictionary t ON ass.tid = t.tid

JOIN

docs ON ass.doc_id = docs.doc_id

LEFT JOIN activity_properties ap ON a.activity_id = ap.activity_id

LEFT JOIN compound_properties cp ON parent.molregno = cp.molregno

LEFT JOIN compound_records cr ON parent.molregno = cr.molregno

LEFT JOIN source src ON cr.src_id = src.src_id

LEFT JOIN compound_structures cs ON parent.molregno = cs.molregno

WHERE md.max_phase = 4 AND a.standard_type IN ('EC50', 'IC50', 'Emax') AND ass.type = 'F';

Table 5: The SQL for extracting the bioassays for a specific drug from ChEMBL

SELECT DISTINCT

parent.chembl_id AS chembl_id,

parent.pref_name AS Name,

md.max_phase,

cr.compound_key AS PubChem_CID,

t.pref_name AS target_names,

ass.assay_type,

ass.description AS assay_info,

a.standard_type,

a.standard_value,

a.standard_units,

ap.type,

ap.relation,

ap.value,

ap.units,

cp.MW_FREEBASE AS MW,

cp.full_mwt AS full_MW,

docs.chembl_id AS document_chembl_id,

docs.journal AS journal,

docs.year AS year,

docs.volume AS volume,

docs.issue AS issue,

docs.first_page AS first_page,

docs.last_page AS last_page,

docs.pubmed_id AS pubmed_id,

docs.doi AS doi

FROM molecule_dictionary md

JOIN molecule_hierarchy mh ON md.molregno = mh.molregno

JOIN molecule_dictionary parent ON mh.parent_molregno = parent.molregno

JOIN activities a ON md.molregno = a.molregno

JOIN assays ass ON a.assay_id = ass.assay_id

JOIN target_dictionary t ON ass.tid = t.tid

JOIN docs ON ass.doc_id = docs.doc_id

LEFT JOIN activity_properties ap ON a.activity_id = ap.activity_id

LEFT JOIN compound_properties cp ON parent.molregno = cp.molregno

LEFT JOIN compound_records cr ON parent.molregno = cr.molregno

LEFT JOIN source src ON cr.src_id = src.src_id

LEFT JOIN compound_structures cs ON parent.molregno = cs.molregno

WHERE md.max_phase = 4 AND a.standard_type IN ('EC50', 'IC50', 'Emax') AND parent.chembl_id = 'xxxxxx';

Table 6: The summary statistics of the Bioassay_Virus_Drug

| Column | Type | Description | Number of records |
| --- | --- | --- | --- |
| chembl_id (K) | CHAR | The unique identifier defined by ChEMBL for a drug | 103193 |
| Name | CHAR | The name of a drug | 103193 |
| max_phase | INT | The phase of the drug development. ChEMBL defines 4 for an approved drug. | 103193 |
| PubChem_CID | CHAR | The unique identifier defined by PubChem for a drug | 103193 |
| target_names | CHAR | The curated target names of the molecular systems being investigated in bioactivity assays | 103193 |
| assay_type | CHAR | The assay was conducted to measure compound bioactivity | 103193 |
| assay_info | CHAR | Description of the assay | 103193 |
| standard_type | CHAR | The activity measurement that has been standardised across different assays | 103193 |
| standard_value | FLOAT | The value of the standardised measurement for the activity type | 99955 |
| standard_units | CHAR | The units associated with the standard_value | 100863 |
| type | CHAR | The original activity measurement type reported in the assay | 4414 |
| relation | CHAR | The relationship between the measured value and the reference | 4414 |
| value | FLOAT | The raw numeric result from the assay corresponding to the activity type | 4414 |
| units | FLOAT | The measurement unit of the value as reported in the assay | 4414 |
| MW | INT | The parent form of the molecule after removing any salts | 103193 |
| full_MW | INT | The molecular weight of the full compound, including any salts or hydrates present | 103193 |
| document_chembl_id | CHAR | The unique identifier defined by ChEMBL for an assay or a literature | 103193 |
| journal | CHAR | The name of the scientific journal where the assay was published | 77893 |
| year | INT | The year of a publication | 102702 |
| volume | CHAR | The volume number of the journal issue | 77556 |
| issue | CHAR | The specific issue number within the volume | 63606 |
| first_page | CHAR | The first page number where the article starts | 77560 |
| last_page | CHAR | The last page number where the article ends | 77560 |
| pubmed_id | CHAR | The identifier for the publication in the PubMed database | 75929 |
| doi | CHAR | The Digital Object Identifier for the publication | 102886 |
| Virus (FK) | CHAR | The abbreviated form of virus name | 103193 |
| Virus.Name (FK) | CHAR | The full name of virus | 103193 |

### S2.4 Tables of PK Parameters

The PK parameters are extracted from the DrugBank Pharmacology sections of each drug. The volume of distribution (Vd), clearance (CL), and protein binding (fu) values are stored in the corresponding subsections for each individual under Pharmacology. The bioavailability (F) and the absorption rate constant (ka) or the time to maximum concentration (Tmax) are stored in the Absorption subsection under Pharmacology.

The general method for extracting PK parameter values can be separated into 1. Rule-based extraction; and 2. LLM-based extraction. The pre- and post-processing steps for these two methods are included in the method details. Later, the combination of PK values from Lombardo et al. (2018) and Mamada et al. (2021) is described.

#### S2.4.1 Clearance and Volume of Distribution

We have applied four steps to curate the PK dataset on clearance and volume of distribution, as shown in Figure S5. We first extract the CL and Vd values using the rule-based method. Second, we combined this dataset with the values from Lombardo et al. (2018) and Mamada et al. (2021). After these two steps, we had covered CL and Vd values for only 1276 drugs. Therefore, we applied GPT-based large-language model (LLM) extraction to obtain the PK values. In the end, we also used the QSPR model to predict CL and Vd values from their SMILES. The outcome of the drug number coverage is shown in each step in Figure S5.

1. For the rule-based methods for clearance and volume of distribution, we first remove all references in the squared brackets and the extra information in the brackets. We also removed the newline character (“\n”) and the comma. We split the string by using the following rule: “was|[*]|is|between|approximately|averaging|averaged|of|about|[,]|and|than|[:]|were” so that we can get the terms before and after these words.

523 drugs

Rule-based extraction

1276 drugs

Combined with publications

1424 drugs

LLM-based extraction

2228 drugs

QSPR-predicted

**Figure S5:** The total drug coverage of clearance and volume of distribution after different methods.

Later, we separate the sentence by spaces and extract the number and units by position. However, if the records contain a range of values such as “2-3 mg/min” or “2±0.5 mg/min”, then we check the second position by using the UTF-8 code. For the “2-3 mg/min”, we will output the 2.5 mg/min as the clearance value. For the “2±0.5 mg/min”, we will extract the 2 mg/min as the clearance value. We will extract the unit from either position 2 (for the normal case, “2 mg/min”) or position 4 (for both cases, “2±0.5 mg/min” and “2-3 mg/min”).

Then, based on the extracted value and units, we convert the value to “L/h” as the standard units for clearance and to “L” as the standard units for volume of distribution. Thus, we have completed the rule-based method for extracting clearance and volume of distribution values with their units.

2. For the second step, we add the chembl_id to Lombardo’s dataset, as they only have the cas_id. Similarly, Mamada’s dataset contains only the drug name. We first capitalize all drug names from Lombardo’s and Mamada’s datasets, so that we can use the soft match and hand-pick to match the drug name to the chembl_id. Later, we convert Lombardo’s units (“mL/min/kg” for CL and “L/kg” for Vd) to our standard units (“L/h” for CL and “L” for Vd). For Mamada’s dataset, we also convert the values to our standard units (“L/hr/kg” to “L/h” for CL and “L/kg” to “L” for Vd). We assume that our standard patient is 70kg. Afterwards, we merge the CL and Vd values from both Lombardo’s and Mamda’s datasets into the PK dataset.
3. For the third step, we want to extract more information from the DrugBank database. We use GPT-based prompting to extract CL and Vd values from each drug's description, as the rule-based method omits several records. For the prompt, we use the

“Can you extract information and create a table with three columns (Parameter, Value, and Unit) to include volume of distribution or Vd or distribution (labeled as Vd); and clearance or CL (labeled as CL) from”

as the prompt to extract the CL and Vd values from the DrugBank Clearance and Volume of Distribution section description. To accelerate extraction, if the records do not contain any numbers or words (e.g., NA), we will skip them. As the response may be glitched and does not extract properly, we would consider adding some examples to the prompt as

“For example, given the statement of ‘0.80 ± 0.24 L/hr/kg [asymptomatic, HIV-1-infected adult patients receiving single (IV dose of 150 mg]’, we extract the row of ‘CL | 0.8 | L/hr/kg |’.”

Therefore, we develop a framework that moves from zero-shot prompting to few-shot prompting, enabling us to improve the performance of GPT prompt engineering.

4. For the fourth step, we use the pre-trained QSPR models to predict the CL and Vd from the SMILES structure. Additionally, we use SQL in Table 7 to extract the SMILES for a given drug chembl_id.

Table 7: The SQL for extracting the SMILES for a specific drug from ChEMBL

SELECT

md.chembl_id,

cs.canonical_smiles AS smiles

FROM

molecule_dictionary md

JOIN

compound_structures cs

ON md.molregno = cs.molregno

WHERE

md.chembl_id = 'xxxxxx';

Then, we loop over the SMILES to predict CL and Vd on a log10 scale with the units of log(mL/min/kg) and log10(L/kg), respectively. Then, the CL and Vd can be recovered to the original scale and converted to the standard units (“L/h” for CL and “L” for Vd). Here, we assume the standard patient weighs 70kg.

Therefore, we store all information from Steps 1 to 3 in the “Drug_PK_Property” table, with a source column indicating whether the data is from DrugBank or publications. The predicted values from Step 4 are stored in the “Drug_PK_Pred” tables.

#### S2.4.2 Protein Binding

The methods for protein binding follow the same procedures as those for volume of distribution and clearance extraction, except for the rule-based extraction. The outcome of each method is shown in Figure S6.

739 drugs

Direct from publications

880 drugs

LLM-based extraction

2228 drugs

QSPR-predicted

**Figure S6:** The total drug coverage of protein binding after different methods.

The main difference happens in the LLM-based extraction. After setting up the original values from Mamada’s dataset, the prompt for extracting the protein binding from DrugBank is

“Can you extract protein binding value from ‘xxx’ with only numbers and without the natural language reply?”

where ‘xxx’ is the description under the Protein Binding section from DrugBank. The value is parsed and stored in the protein binding column. Once we retrieved the columns from DrugBank, we merged the protein-binding values into the “Drug_PK_Property” table. Also, the predicted protein binding is stored in the “Drug_PK_Pred” table.

#### S2.4.3 Bioavailability and Absorption Rate Constant

In DrugBank, the information on bioavailability (F) and absorption rate constant (ka) is stored in the Absorption section of each drug. We use different methods to extract the values of F and ka. The numbers of F and ka are shown in Figures S7 and S8, respectively.

139 drugs

LLM-based extraction

**Figure S7:** The total drug coverage of bioavailability using LLM.

258 drugs

Rule-based extraction + pharmacokinetic processing

**Figure S8:** The total drug coverage of the absorption rate constant using the rule-based method and postprocessing.

- To extract the values of F, we use the following prompts to extract the values:

“Can you extract information and create a table with 3 columns (Parameter, Value, and Unit) to include Tmax; absorption rate constant (labeled as Ka); and Bioavailability from ‘xxx’ in a table and without the natural language reply?”

As stated from the prompt, we also extract the Tmax or ka. However, after checking the Tmax and ka values, we found that Tmax or ka is affected by severe hallucination. We decided to abandon the Tmax or ka extraction. Instead, we will use the rule-based method to extract Tmax or ka.

Following the bioavailability method, we extract the value and convert it to a percentage. Later, we store the bioavailability into the “Drug_PK_Other” tables.

- For the extraction of ka, we use the following rules first to clean the absorption description. We first remove the square brackets and their contents. Later, we extract both tmax and ka from the following rules:

“(ka)[^0-9]*([0-9]+\\.?[0-9]*(?:e[+-]?[0-9]+)?)(?:\\s*to\\s*([0-9]+\\.?[0-9]*))?\\s*([a-zA-Z/]+)?” and

“(tmax)[^0-9]*([0-9]+\\.?[0-9]*(?:e[+-]?[0-9]+)?)(?:\\s*(?:to|-)\\s*([0-9]+\\.?[0-9]*))?\\s*([a-zA-Z/]+)?”.

These two rules extract patterns after a “ka” or “Tmax” that contain a number before the decimal point and another after. At the end of the pattern, we also extract the words right after the decimal number. Sometimes the Tmax value is hidden in the Cmax sentence. For example, the Tmax value can come after “The maximum concentration (Cmax) can be achieved after 1.7 hours.” Therefore, we also extract the “Cmax” pattern using the same rule. Afterwards, we calculate the extraction using the first-order absorption assumption and a single-dose input

Where .

Based on this calculation, we can solve ka in terms of Tmax, CL, and Vd. However, there are two solutions to this equation. We can examine the solutions by applying back to the Tmax function and compare the calculated Tmax with the extracted Tmax value. Therefore, we can get the ka values from the extracted Tmax values. We added the Tmax column to the “Drug_PK_Property” table so we can align Tmax with CL and Vd for each drug. Using the calculation stated before, we can gather the ka column in the “Drug_PK_Property” table.

Once we get the “Drug_PK_Other” and “Drug_PK_Property” tables, we integrate the “Drug_PK_Other” table to the “Drug_PK_Property”. Therefore, we only show the table details of the “Drug_PK_Property”. Because the QSPR model does not predict absorption, the bioavailability and absorption rate constant are not included in the “Drug_PK_Pred” table.

In summary, the PK tables include the summary statistics for “Drug_PK_Property” and “Drug_PK_Pred” in Tables 8 and 9, respectively. In these two tables, the number of records is the total count of each PK parameter, which may contain duplicates. The unique drug number is referenced in Figures S5 to S8.

Table 8: The summary statistics of the Drug_PK_Property

| Column | Type | Description | Number of records |
| --- | --- | --- | --- |
| DrugBank_id (K) | CHAR | The unique identifier defined by DrugBank for a drug | 1856 |
| Name | CHAR | The name of a drug | 1856 |
| chembl_id (FK) | CHAR | The unique identifier defined by ChEMBL for a drug | 1683 |
| volume_of_distribution | FLOAT | The value of the volume of distribution for a drug | 1856 |
| vd_unit | CHAR | The unit of volume of distribution | 1856 |
| clearance | FLOAT | The value of the clearance for a drug | 1856 |
| cl_unit | CHAR | The unit of clearance | 1856 |
| Rb | FLOAT | The value of retinoblastoma protein binding for a drug | 818 |
| Source | CHAR | The origin of the PK information, i.e. DrugBank, Lombardo et al. or Mamada et al. | 1856 |
| doi | CHAR | The Digital Object Identifier for the publication | 1856 |
| bioavailability | INT | The value of bioavailability for a drug | 83 |
| keyword | CHAR | The entity name of Tmax or Cmax | 367 |
| tmax | FLOAT | The value of the Tmax for a drug | 367 |
| unit | CHAR | The unit of Tmax | 367 |
| ka | FLOAT | The value of the absorption rate constant for a drug | 369 |
| ka_unit | CHAR | The unit of absorption rate constant | 1856 |

Table 9: The summary statistics of the Drug_PK_Pred

| Column | Type | Description | Number of records |
| --- | --- | --- | --- |
| chembl_id (K, FK) | CHAR | The unique identifier defined by ChEMBL for a drug | 2228 |
| Name | CHAR | The name of a drug | 2228 |
| smiles | CHAR | The SMILES structure of a drug | 2228 |
| Rb | FLOAT | The predicted value of retinoblastoma protein binding for a drug | 2228 |
| volume_of_distribution | FLOAT | The predicted value of the volume of distribution for a drug | 2228 |
| clearance | FLOAT | The predicted value of the clearance for a drug | 2228 |

### S2.5 Drug Tissue Partition

For the “Drug_Tissue_Partition” table, we assume the drug partition is well perfused. Furthermore, we assume that the rate at which a drug enters a tissue depends on the rate of blood flow to that tissue, the amount of tissue volume, and the rate at which the drug moves between blood and tissue.

Based on instant equilibria, we would require the pH of the tissue and pKa and pKb of the drug (Rodgers et al., 2005). Furthermore, we would also require the fractional tissue volumes of plasma and blood cells (Rodgers & Rowland, 2007). For the organ-level physiological data, we collected pH and tissue composition from Ye et al. (2016) and Schmitt (2008).

Along with the pKa, pKb, logP and logD information from the “Approved_Drugs” and the blood-to-plasma concentration and the drug fraction from Mamada et al. (2021), we can start to calculate the tissue partition coefficient for each drug using the formula from Rodgers et al. (2005) and Rodgers & Rowland (2007). Specifically, if we lack the information on protein binding for a specific drug, we would assume the free drug fraction is 1.0 and the blood-to-plasma concentration is 0.7. Otherwise, we will use the blood-to-plasma concentration and the drug fraction from Mamada et al. (2021).

In summary, we obtained partition coefficients across 11 organs for 715 drugs in the “Drug_Tissue_Partition” table. The detail of this table is listed in Table 10.

Table 10: The summary statistics of the Drug_Tissue_Partition

| Column | Type | Description | Number of records |
| --- | --- | --- | --- |
| chembl_id (K, FK) | CHAR | The unique identifier defined by ChEMBL for a drug | 7865 |
| Tissue | CHAR | The specific biological tissues or sites | 7865 |
| Neutral.lipids | FLOAT | The lipid fraction in tissues that are neutral lipids | 7865 |
| Neutral.phospholipids | FLOAT | The tissue phospholipids that are electrically neutral at physiological pH | 7865 |
| Intracellular.water | FLOAT | The water content inside tissue cells | 7865 |
| Extracellular.water | FLOAT | The water content outside tissue cells | 7865 |
| Tissue.conc.of.acidic.  phospholipids | FLOAT | The concentration of acidic phospholipids in a tissue | 7865 |
| Albumin.ratio | FLOAT | The ratio of drug bound or partitioned into albumin | 7865 |
| Lipoprotein.ratio | FLOAT | The distribution ratio of a drug into lipoproteins | 7865 |
| pH | FLOAT | The measure of acidity or alkalinity of the environment where partitioning occurs | 7865 |
| Partition.coefficient | FLOAT | The ratio of concentrations of a drug partitioning in a tissue | 7865 |

### S2.6 Dosing Regimen

For the Dosing Regimen, we extract the tables from the following websites: DailyMed (https://dailymed.nlm.nih.gov/dailymed/), openFDA (https://open.fda.gov/), Drugs.com (https://www.drugs.com/), Medscape (https://reference.medscape.com/drugs/antimicrobials), and Farmacotherapeutisch Kompas (https://www.farmacotherapeutischkompas.nl/). Specifically, we use the last two websites (Medscape and Farmacotherapeutisch Kompas) for our curation of antiviral drugs, as the Antimicrobials page on Medscape lists them.

Dosing information was compiled from DailyMed, openFDA, Drugs.com, and additional private datasets; full extraction details will be provided upon publication.

1. We first clean the administration route information and create a new column indicating whether the drug belongs to Oral, IV Bolus or IV Infusion.
2. Then, we extract the loading dose, maintenance dose, dose interval, duration and maximum dose from the table. We extract the loading dose and maintenance dose from the corresponding columns by directly reading the values and converting them to mg. We also extract the dosing interval from the regimen column; e.g., “BID” indicates twice a day, and the dosing interval is set to 12 hours. Also, the duration of using drugs is converted to days.
3. Afterwards, we go through the whole list of dosing regimens, and convert every value to the standard units, which are “mg” for the dose information (loading, maintenance and maximum), “h” (hour) for the dosing interval, and “d” (day) for the duration.
4. Once we got the extracted tables, we curated extra dosing regimens and maximum dose from Medscape and Farmacotherapeutisch Kompas.

Therefore, we gather the dosing regimen information for the "Dosing_Regimen_Drug" table. In total, we have gathered 371 drug dosing regimens. Furthermore, we simulate Cmax and AUC based on the dosing regimen records, enabling the user to check toxicity from a modified input. The summary of the table is listed in Table 11. Specifically, the Cmax and AUC information of maximum dosing regimens is limited due to the PK parameters.

Table 11: The summary statistics of the Dosing_Regimen_Drug

| Column | Type | Description | Number of records |
| --- | --- | --- | --- |
| Name | CHAR | The name of a drug | 573 |
| chembl_id (K, FK) | CHAR | The unique identifier defined by ChEMBL for a drug | 573 |
| group_id | CHAR | The unique identifier for the treatment group in a clinical trial record | 573 |
| Administration.Route | CHAR | The original record of the administration route of a drug | 573 |
| AR | CHAR | The clean administration route for a drug, i.e. Oral, IV Bolus and IV Infusion | 573 |
| Common.Formulations | CHAR | The formulation of a drug | 573 |
| FDA.Source | CHAR | The source of the clinical trial record or the dosing regimen record | 573 |
| load.dose | FLOAT | The amount of loading dose | 573 |
| load.unit | CHAR | The unit of loading dose, measured in mg | 573 |
| main.dose | FLOAT | The amount of maintenance dose | 573 |
| main.unit | CHAR | The unit of maintenance dose, measured in mg | 573 |
| dose.int | FLOAT | The dose interval, measured in hours | 573 |
| duration | FLOAT | The duration of a dosing regimen | 573 |
| dura.unit | CHAR | The unit of duration, measured in days | 573 |
| max.dose | FLOAT | The maximum amount of accumulated dose per day | 573 |
| max.unit | CHAR | The unit of maximum dose, measured in mg | 573 |
| inf.duration | FLOAT | The infusion duration | 573 |
| inf.unit | CHAR | The unit of infusion duration, measured in minutes | 573 |
| max.Cmax | FLOAT | The Cmax is calculated from the dosing regimen with the highest loading and maintenance dose within the daily maximum dose | 131 |
| max.AUC | FLOAT | The AUC is calculated from the dosing regimen with the highest loading and maintenance dose within the daily maximum dose | 131 |
| max.Cmax.unit | CHAR | The unit of the maximum Cmax, measured in mg/L | 573 |

### S2.7 Drug Model Library

For the Drug Model Library, we mainly want to connect the model folders to the “Approved_Drugs” table. Therefore, we generated the table and added the chembl_id column. Therefore, we extract the model name from the PopPK model file name and obtain information from the model metadata and model settings. The summary of the “Drug_Model_Library” is listed in Table 12. In total, we curated 19 models for the drugs, and 14 of them belong to the approved drug list, and 5 of them are for the other phase drugs.

Table 12: The summary statistics of the Drug_Model_Library

| Column | Type | Description | Number of records |
| --- | --- | --- | --- |
| drug | CHAR | The name of a drug | 19 |
| is.prodrug | BOOL | Is the drug a prodrug or not | 19 |
| n.cmt | INT | The number of compartments defined in the PopPK model | 19 |
| is.covariate | BOOL | Is any covariate defined in the PopPK model | 19 |
| population | CHAR | The description of the population defined in the PopPK model | 19 |
| route | CHAR | The administration route defined in the PopPK model | 19 |
| author | CHAR | The author of the publication | 19 |
| year | CHAR | The year of the publication | 19 |
| DOI | CHAR | The Digital Object Identifier for the publication | 19 |
| journal | CHAR | The name of the scientific journal where the PopPK model was published | 19 |
| description | CHAR | The one-liner of the PopPK model description | 19 |
| chembl_id (K, FK) | CHAR | The unique identifier defined by ChEMBL for a drug | 14 |
| file_name | CHAR | The file name of the PopPK model | 19 |
